# Evaluating trends in new cases of Work-Related Mental Ill-health after introduction of The Health and Safety Executives’ Management Standards

**DOI:** 10.64898/2026.05.08.26352738

**Authors:** M. Gittins, Z. Iheozor-Ejiofor, M. Carder, AM. Money, I. Iskandar, N. Gartland, D. Fishwick, M. Seed, G. McHale, L. Byrne, M. van Tongeren

## Abstract

**Background:** Work-related stress (WRS) accounts for 52% of self-reported work-related ill-health. In 2004, the Health and Safety Executive (HSE) developed the Management Standards (MS), aimed at helping organisations reduce WRS. This work investigates WRS post implementation, with reference to six MS risk factor domains: *control, support, relationships, demand, change, and role*.

**Method:** Cases of WRS were extracted from The Health and Occupation Research (THOR) database and mapped to the six domains. Trends in WRS incidence rates attributed to each of the domains were split at 2004 and compared with the overall WRS trend using mixed generalised regression models.

**Results:** Before 2004, annual incidence in WRS increased by 1.4%(-0.5%,3.1%), whereas after 2004, there was a decrease of -0.9%(-1.5%,-0.2%), based on 10,815 WRS cases reported between 1996 and 2019. Three of the six MS domains (*demands, relationships, and change*) were reported in ∼82% of cases. Pre-2004, four of the six domains were observed to be increasing per year. Post 2004, cases increasingly contained multiple precipitating events e.g. *demands + another* (+2.6% per year) and *relationships + another (6*.*1%)*. Reports of the two most common domains decreased post 2004 (*demands* -0.46%, *relationships* -0.55% per year), whereas incidence in less common domains increased (change 1.1%, support 2.4%, control 4.8%, role 4.7%).

**Conclusion:** Trends in WRS, and their common risk factors appear to be decreasing gradually, since introduction of MS in 2004. However, less common risk factors are becoming more prominent, contributing to reporting of WRS with multiple risk factors.

## Introduction

Mental ill-health problems have overtaken musculoskeletal disorders as the main cause of self-reported work-related ill-health (WRIH) in the UK. Work-related stress, anxiety and depression accounted for 56% of new cases of WRIH in the financial year 2024/25.^1^ The burden of work-related mental ill-health (WRMIH) problems remains high, with cases of stress, anxiety and depression accounting for 22.1 million working days lost in 2024/25. New cases of WRMIH were estimated to account for £5.2 billion per year in 2013/14, 55% of the total cost of WRIH.^2^ In 2024/25 this estimate has risen to £9.1 billion per year.^1^

Work-related stress has been made a prioritisation area, and the Health and Safety Executive (HSE) has devised a strategy to reduce it.^3 4^ Specifically, HSE identified the potential for stress at work to be risk assessed, hence managed like other hazards in the workplace,^5^ and launched the Management Standards (MS) in 2004.^6^

The MS were intended to be applied at an organisational, rather than individual, level. This approach identified six areas that, if incorrectly managed, could be associated with increased sickness absence, poor health and well-being and lower productivity amongst workers.^7^ These six areas or domains were: demands, role, change, control, support and relationships. There was an expectation that an organisation would regularly conduct stress risk assessments and take sufficient action to tackle any problems identified through risk assessment. HSE implementation began in 2004^2^ and additionally produced a range of detailed guidance on how to use the MS risk assessments and indicator toolkits.^8^

The MS are now a commonly used tool for managing and reducing work-related stress already present within a work-force and thought related to occupational factors.^9 10 11^ The potential utility of the MS in reducing WRS was previously supported by work in a large community-based Health and Social Services Trust.^7^ Here the HSE Stress Indicator Tool responses from within a cross-sectional study of workers were positively associated with job satisfaction and negatively associated with job-related anxiety and job-related depression^7 12^. However, little evidence currently exists of the influence the MS have had on change in new work-related stress (WRS) cases rather than in prevalent cases. In-house HSE evaluations of MS, concluded that the approach “works well in principle but less so in practice”^13^ as “organisations experience problems following through and implementing risk reduction interventions” ^13^ The HSE’s own trends in new and long-standing cases using self-reported WRS have remained broadly flat between 1998 and 2020 showing very little fluctuation.^14^ Self-reported WRS cases are dependent on a person’s own belief that their work has caused their stress.^15^ In contrast, The Health and Occupation Research (THOR) network^16^ collects incident cases of WRS which have been assessed by an occupational health (OH) physician to be related to or caused by the work of the patient.

Here we model the trends in new cases of WRS as reported by OH physicians to the THOR scheme in the UK and compare trends before and after implementation of the MS in 2004. We explored WRS trends overall and WRS trends associated with the six MS domains.

## Methods

The methodology of THOR has been previously documented.^16^ Briefly, new cases of WRIH are reported by either ‘core’ reporters, who report monthly throughout the year, or ‘sample’ reporters who report for one randomly selected month per year. For each case, diagnosis, age, sex, first half of postcode, industry, occupation, causal agent(s) (in the cases of mental ill-health, precipitating events), and date of symptom onset are recorded. Occupation and workplace are subsequently coded using Standard Occupational Classification and Standard Industrial Classification,^17 18^ respectively. Diagnosis is coded to ICD-10 ^19^ whilst the agents / precipitating events are coded using in-house coding systems.

Cases of WRMIH as reported by OPs to the Occupational Physicians Reporting Activity (OPRA; data available for between 1996-2019) were included. Due to the influence of COVID-19 on reporting and its potential influence on trends of WRIH, data after 2020 were excluded here. The ICD-10 code MIH diagnoses broadly map to anxiety/depression; post-traumatic stress disorder; work-related stress; alcohol or drug abuse; psychotic episode; other psychiatric problems. Cases diagnosed as ‘work-related stress’, ‘anxiety/depression’ and certain ‘other psychiatric problems’ (e.g. adjustment disorder or stress-like symptoms) based on their ICD-10 coding for MIH were included; for the purpose of this analysis this selection will be referred to as work-related stress (WRS).

For each case reported to OPRA, a maximum of three precipitating events may be reported by the OP, which are in turn mapped to THOR’s in-house coding system (see Table 1). In most cases mapping was straightforward but one in-house code, ‘organisational factors’, could potentially be mapped to any of the three MS: *control, support* or *role*. The reported descriptive precipitating events of cases not easily mapped were independently reviewed and mapped to the relevant MS domain(s) by two project assistants with any discrepancies reconciled by the project lead.

**Table 1:** Mapping of in-house codes to HSE management standards.

| **In-house classification** | **HSE management standards** |
| --- | --- |
| 1.1 workload, over demand, targets: 1.2 work schedule, hours: 1.3 travel, working patterns: 1.4 isolation: 1.6 under load, boredom: 1.7 responsibilities: 6.1 physical conditions: 6.2 environmental factors | Demands |
| 3.0 interpersonal relationships: 4.0 inequality and unfair treatment in the workplace | Relationships |
| 2.1 Change of management, organisational change: 2.2 changes in ways of work inc. technological change: 2.3 change of work content: 2.4 reduction in resources inc. staff: 2.5 relocation | Change |
| 1.5 Organisational factors – including role poorly defined, low job control, lack of management support and poor management | Control – after detailed review |
|  | Support – after detailed review |
|  | Role – after detailed review |

We initially report an overall trends analysis, as in the trend associated with all WRS reported to the scheme. Then the MS domain(s) assigned to a WRS case were categorised in two ways; the first based on the `combinations’ of (up to three) precipitating events mapped to a MS domain, and the second where there was `any mention’ of the individual MS domain as a precipitating event regardless of whether other MS domains are mentioned. For example, a reported case of WRS may have two precipitating events one mapped to *control* and one to *support*. This case would then be allocated to: *control & support* in combination; *any control*; *any support*. When all cases were aggregated and combinations of domains were collated, fifty-three unique combinations were observed, of which 79% cases were represented by just four unique combinations, and 43 of the 53 each contained <1% WRS cases (see Supplementary Material Table S1 for full list). Under review by the study team, we reclassified these 53 groups into more succinct common combinations, defined as: `Domain unknown’, `Demands only’, `Relationships only’, `Change only’, `Support only’, `Demands + any other’, `Relationships + any other’, and `Any other combination’.

To investigate the trends in WRS, STATA v17 was used to fit a set of multilevel mixed effect negative binomial models with a random intercept. The random effects intercept term adjusted for within and between reporter correlation due to reporter clustering. This method allows for variation in the number of reports between physicians (for example, core versus sample and due to non-response).^20^ To describe the trends in WRS, calendar year was represented in the model as a single continuous linear variable (mean centred), and separately as two linear splines split at 2004 (i.e. the year MS were introduced). The trends analysis was carried out for all WRS cases and then repeated separately within each of the `common combinations’ (e.g. those cases for *demands only* and *demands + any other*) and separately for `any’ report of each MS domains (e.g. *any control*; *any support*, etc). Additionally, for descriptive purposes each model was also fitted with calendar year as a categorical variable. The subsequent Relative Risks for each calendar year against the reference year 2019, were plotted in order to describe the trends over time. The calendar year effect was further adjusted for a set of potential confounders commonly used in THOR studies; reporter type, first month as a reporter and calendar month of report, and annual average working population as offset adjustment.

An awareness campaign occurred for two years post 2004 implementation, meaning effectiveness may have been delayed. A sensitivity analysis assessed the assumption implementation in 2006 was the key linear split point.

## Results

Between 1996-2019, 10,815 eligible WRS cases were identified based on reported diagnostic category i.e. ‘work-related stress’ (∼50%), ‘anxiety/depression’ (∼56%) and ‘other psychiatric problems’ (3%). Of which 8,201 (76%) WRS were mapped to the MS, leaving 2614 that could not be assigned. Of the 8,201 eligible cases mapped to the MS, 61% were female and the mean age was 45 years (oldest at ∼75 years). The industry sectors with the highest proportion of cases were health and social care (44%), public administration and defence (15%), and education (14%). The most frequently reported occupations were nurses (10%), teachers (5%), doctors (3%) and administration/clerical officers (3%).

Tables 2 & 3 describe the WRS cases by MS domain under our two categorisation definitions i.e. any mention of each MS domain and the identified common combinations. The MS domains, *demands*, and *relationships*, respectively were reported most often at 35% and 30% of all cases with any mention. Additionally, 24% of cases were mapped to *only demands* and 24% to *only relationships* as contributory factors i.e. no other additional factor was reported. Of the 14.8% of cases with more than one domain reported (see Table S1) *demands + another domain* made up 67%, i.e. 10% of all cases. Further descriptive statistics can be found in the supplementary material. A full description of count of number of cases reported per month (i.e. the basis for the trends model) by MS domains can be found in the Supplementary Table S2. Figure S3 reports stacked counts and percentages of new cases of WRS split by the common aggregated domains. The stacked percentages for those cases not assigned a MS domain have decreased as a proportion of responses over time, whereas multi-combinations such as *demands+other* and to a lesser extent *relationships+other* have increased as a proportion of WRS cases.

**Table 2:** Annual percentage change (95% Confidence intervals) in Work Related Stress (WRS) trends for total WRS and their associated common combinations of Management Standards (MS) terms mentioned: OPRA data (1996-2019) subsequently split before and after MS implementation in 2004.

| **Trends (1996-2019)** | **Total WRS** | **Not-MS Assigned** | **Demands Only** | **Relationships Only** | **Change Only** | **Support Only** | **Demands + Other** | **Relationships + Other** | **Any Other Combinations** |
| --- | --- | --- | --- | --- | --- | --- | --- | --- | --- |
| Calendar Yr (Mean Centred)<sup>a</sup> | -0.43<br>(-0.95,0.09) | -3.03<br>(-4.10,-1.94) | -0.99<br>(-2.04,0.07) | 0.48<br>(-0.58,1.55) | -1.34<br>(-3.13,0.49) | -0.88<br>(-3.36,1.67) | 3.11<br>(1.45,4.79) | 7.26<br>(4.10,10.51) | 0.73<br>(-2.03,3.58) |
| Random Effect Variance | 0.13 | 0.39 | 0.33 | 0.37 | 0.53 | 0.99 | 0.84 | 1.34 | 0.87 |
| AIC | 15125.9 | 8563.5 | 8519.5 | 8279.5 | 4287.7 | 2677.0 | 5053.4 | 1812.0 | 2386.2 |
| No cases (%) | 10,815 | 2614(24.2) | 2570(23.8) | 2546(23.5) | 821(7.59) | 439(4.06) | 1,180(10.9) | 290(2.68) | 355(3.28) |
| **Trends (split at 2004)** |  |  |  |  |  |  |  |  |  |
| Calendar Yr(pre2004)<sup>a</sup> | 1.36<br>(-0.47,3.12) | -5.69<br>(-8.76,-2.51) | 4.15<br>(0.48,7.95) | 12.30<br>(7.89,16.88) | -2.84<br>(-8.33,2.99) | 15.9<br>(4.85,28.1) | 5.76<br>(-0.49,12.39) | 16.6<br>(-0.18,36.2) | -2.75<br>(-11.75,7.17) |
| Calendar Yr(2004-)<sup>a</sup> | -0.86<br>(-1.50,-0.21) | -1.29<br>(-2.72,0.15) | -2.17<br>(-3.47,-0.85) | -1.72<br>(-2.99,-0.44) | -0.94<br>(-3.24,1.42) | -3.81<br>(-6.77,-0.76) | 2.61<br>(0.61,4.65) | 6.14<br>(2.46,9.94) | 1.53<br>(-1.94,5.12) |
| Random Effect Variance | 0.13 | 0.39 | 0.33 | 0.38 | 0.54 | 0.97 | 0.83 | 1.34 | 0.88 |
| AIC | 15123.3 | 8558.7 | 8513.0 | 8247.6 | 4289.4 | 2668.0 | 5054.7 | 1812.7 | 2387.7 |
| N cases pre2004 (%) | 2106 | 828(39.3) | 496(23.6) | 344(16.3) | 171(8.1) | 47(2.2) | 145(6.9) | 20(0.95) | 55(2.61) |
| N cases post2006(%) | 8709 | 1786(20.5) | 2074(23.8) | 2202(25.3) | 650(7.5) | 392(4.5) | 1035(11.9) | 270(3.1) | 300(3.44) |

a = Adjusted for calendar month, first month as reporter, and 'core' vs 'sample' reporter

The percentage change in Incidence Rate Ratios (IRR) and 95% Confidence Intervals (95% C.I.) per year in the annual trends are reported in Table 2 and 3. Trends in new cases of WRS between 1996 and 2019 (see Table 2) were -0.43% (-0.95%,0.09%) per year. Between 1996 and 2004, WRS increased gradually at 1.4% (-0.5%, 3.1%) per year, whereas for 2004-2019, a -0.86% (-1.5%, -0.9%) decrease per year was observed. Figure 1 describes the incidence rate per year (compared to 2004) in WRS. For not-assigned cases, a 3% (-4.1%, -1.9%) decrease for full 1996-2019, translated into -5.7% (-8.8%, -2.5%) and -1.3% (-2.7%, 0.2%) pre-and post-2004. Table 2 also reports trends associated with monthly counts of the common combinations of domains, whereas Table 3 reports trends associated with `any’ mention of a domain.

**Table 3:** Annual percentage change (95% Confidence intervals) in trends of Work-Related Stress (WRS) due to ‘any’ mention of Management Standards(MS): Trends for complete study period (OPRA data 1996-2019) and split before and after MS implementation in 2004.

| **Trends (1996-2019)** | **Total WRS** | **Not-MS Assigned** | **Demands** | **Relationships** | **Change** | **Support** | **Control** | **Role** |
| --- | --- | --- | --- | --- | --- | --- | --- | --- |
| Calendar Yr(Mean Centred)<sup>a</sup> | -0.43<br>(-0.95,0.09) | -3.03<br>(-4.10,-1.94) | 0.38<br>(-0.50,1.27) | 1.50<br>(0.54,2.48) | 0.48<br>(-0.93,1.91) | 3.84<br>(2.02,5.69) | 2.41<br>(-0.48,5.38) | 5.71<br>(2.43,9.09) |
| Random Effect Variance | 0.13 | 0.39 | 0.25 | 0.35 | 0.51 | 0.74 | 1.35 | 1.21 |
| AIC | 15125.9 | 8563.5 | 10422.2 | 9288.1 | 6203.6 | 4665.6 | 2366.4 | 1839.8 |
| No cases (%)<sup>b</sup> | 10,815 | 2614(24.2) | 3,750(34.7) | 3,218(29.8) | 1,517(14.0) | 1,013(9.4) | 383(3.5) | 263(2.4) |
| **Trends (split at 2004)** |  |  |  |  |  |  |  |  |
| Calendar Yr(pre2004)<sup>a</sup> | 1.36<br>(-0.47,3.12) | -5.69<br>(-8.76,-2.51) | 4.22<br>(1.06,7.47) | 12.9<br>(8.84,17.2) | -2.40<br>(-6.86,2.27) | 13.2<br>(4.97,22.15) | -7.75<br>(-16.5,1.90) | 11.9<br>(-2.14,28.1) |
| Calendar Yr(2004-)<sup>a</sup> | -0.86<br>(-1.50,-0.21) | -1.29<br>(-2.72,0.15) | -0.46<br>(-1.55,0.64) | -0.54<br>(-1.69,0.63) | 1.16<br>(-0.60,2.94) | 2.36<br>(0.19,4.57) | 4.79<br>(1.14,8.58) | 4.72<br>(0.82,8.76) |
| Random Effect Variance | 0.13 | 0.39 | 0.25 | 0.36 | 0.51 | 0.74 | 1.34 | 1.20 |
| AIC | 15123.3 | 8558.7 | 10417.8 | 9253.8 | 6204.0 | 4662.0 | 2364.0 | 1841.0 |
| N cases pre2004 (%)<sup>b</sup> | 2106 | 828(39.3) | 641(30.4) | 402(19.1) | 257(12.2) | 85(4.04) | 55(2.61) | 22(1.04) |
| N cases post2006(%)<sup>b</sup> | 8709 | 1786(20.5) | 3109(35.7) | 2816(32.33) | 1260(14.4) | 928(10.7) | 328(3.8) | 241(2.8) |

a = Adjusted for calendar month, first month as reporter, and ‘core’ vs ‘sample’ reporter AIC = Akaike Information Criteria for within WRS outcome comparison
b = Note cases may have multiple precipitating events hence percentages here may not aggregate to 100%

**Figure 1:**
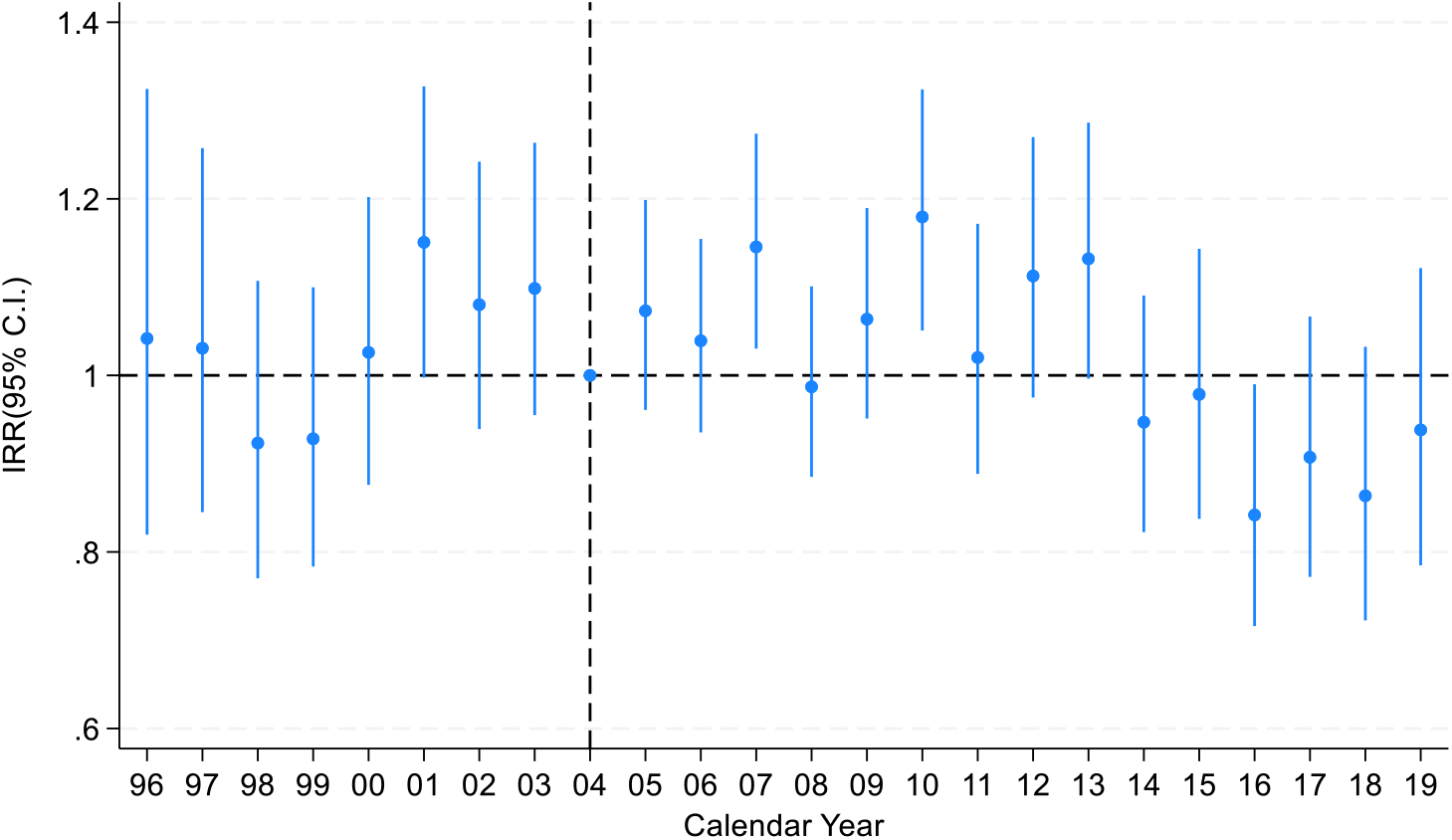
Annual Incidence Rate Ratios IRR (95% Confidence Intervals) for total Work-Related Stress (Management standards implemented in 2004) referenced to 2004.

Within the four most common aggregated domains (Table 2) *demand only, relationships only, change only*, and *support only* (i.e. not reported in combination with another domain) there was little overall change in incidence observed between 1996 and 2019. Whereas prior to 2004 these were increasing at 4%, 12%, 16% per year, respectively, there was a post-2004 gradual decline (-2.1%, -1.7%, -0.94%, and -3.8% respectively). The proportion of cases with multiple precipitating events compared to a single reported event, has increased consistently from 18% in 1996 to 50% in 2019. Two of the commonly observed combinations, *demand + other*, and *relationships + other*, increased between 1996 and 2019, with the rate of increase becoming more gradual post-2004. Supplementary Figure S1 describes these changes in the Incidence Rate per year (compared to 2004) in WRS for each aggregated combined domains individually.

Within the trends analysis for *any mention* of each domain (Table 3) a WRS case may be attributed to multiple *any domains*. Within all 6 domains the percentage change in *any mention* was observed to gradually increase (1996-2019), with strong increases seen in *support* or *role* at 3.8% (2.0%,5.7%), and 5.7% (2.4%,9.1%), respectively. Prior to 2004 there was a 13% and 12% increase in *support* and *role* respectively, compared to 2.3% and 4.7% post-2004. Supplementary Figure S2 describes changes in the incidence rate per year (compared to 2019) in WRS for each domain as defined as any reporting.

We observed no improvement in goodness of fit measures (AIC) of change in calendar trends before or after the 2006 cut-off (Supplementary material sensitivity analysis Table S3).

## Discussion

Previous research concerning the utility of the MS has largely been based on their construct and organisational implementation. Little is currently available to evaluate the introduction of the MS stress indicator tool on the rate of new cases of WRS. This study reported that *demands* and *relationships* were the two most common contributing domain factors assigned to new cases of WRS. Trends in cases where *demands* and *relationships* factors were present have remained largely flat since the MS introduction in 2004, with THOR data suggesting trends were increasing for these two factors prior to 2004. Cases where the two most common domains, *demands* or *relationships*, are the only precipitating event appear to be gradually decreasing or consistent over time whereas more complex cases, i.e. with two or more contributing domains, appear to be increasing. Additionally, *any reports* of the less common domains *change, support, control and role* are increasing post-2004, possibly driven by further reflection of either more complex cases, or more detailed reporting driven by increased awareness.

Several studies have evaluated the MS and health outcomes in a cross-sectional context, some of which sensibly questioned the general approach of the MS and their effectiveness.^9 12 21-26^ Whilst these support the construct of the MS tool as mapping appropriately to health outcomes, they do not comment on whether organisational use of the MS approach had led to a reduction in WRS over time. A Delphi exercise^13^ carried out five years after their introduction questioned whether they work well in practice, as opposed to in principle. In 2010, a HSE psychosocial working conditions survey was undertaken, which showed a decline in psychosocial working conditions, although an upward trend was seen in change and support, and reported conditions were generally positive. That report also identified difficulties with interpretation of trends, including the time lag between organisations first implementing the process and benefits being realised as well as responders’ perception biases. Labour force survey data^27^ relating to trends in anxiety, depression and stress self-reported to be linked to work also show no fall over time (or over the time periods considered specifically in this analysis). If anything, incidence of these cases have been rising since the early 2000s.

Our approach here was different; we aimed to identify if a signal could be picked up in relation to potential change in the incidence of diagnosed cases of WRS (i.e. the overall trends analysis of WRS), at a working population level using data from the national ill-health reporting scheme. Overall, a small downward trend in new cases of WRS was seen, post-2004 introduction of the MS, as opposed to the increase prior to 2004.

Trends in cases where *demands* or *relationships* are the only precipitating events appear to be decreasing, whereas more complex cases with two or more precipitating events associated with different domains appear to be increasing. This may reflect an increase in multi-factorial causes being experienced by those developing WRS in their work-life i.e. cases are simply more complex due to increased presence of secondary additional factors relating to the domains *change, support, control*, and *role*. Alternatively, there may have been an increase in awareness of common precipitating events, due to improved understanding of mental health through campaigns and guidance such as the HSE’s MS, i.e. cases were always complex, but physicians and patients are now better equipped to identify the precipitating events.

The study has several strengths. These cases have been diagnosed by an OH care professional, which will improve the accuracy of the diagnosed health outcome of interest, a likely improvement on case definition based on self-reported WRS. Second, we were able to compare the periods before and after the introduction of the MS, thus providing a suitable period against which to compare case incidence.

Mapping cases *post hoc* into domains within the MS were limited due to difficulties directly matching all the precipitating events with the system developed in-house. For cases without a direct match, we considered and attempted to map directly the original description of precipitating events. Due to varying levels of information reported, there may have been misclassification present. This may have occurred if errors were present in mapping to the original in-house coding system meaning they are either wrongly mapped or not mapped at all to the MS domains, or mapped to the in-house code not directly related to the MS domains (see Table 1). Trends indicate the rate of unassigned cases is decreasing post2004. This may occur, for example, if changes in awareness and understanding of work factors and WRS are reflected in reporting by OPs to the scheme. The Health Management standards themselves may have contributed to these changes raising awareness of particularly work-related stressors. This difficulty in mapping, may in turn influence the trends within individual domains if the available information and subsequent misclassification differs over calendar time.

Precipitating events, and symptoms, of the individual cases are recorded by the OH professional and may not reflect those dynamically experienced by the patient. Thus, here the precipitating events are most important when reporting the case, something which may also change over 20 years due to external factors e.g. changes in training methods, awareness, personal interest. The nature of the workplaces, the proportion of high-risk working population for WRS, and differential coverage of OH provision may have also altered over time though previous work by THOR estimated that it was stable at ∼34% during the 2000s.^28^ Ultimately, the total coverage and change over time in total coverage of active MS implementation within workplaces to monitor and reduce WRS was unknown which may be contributing to these results.

These results may apply differently across industries and occupations, with implementation and impact of MS thought to vary across different industries and occupations.^9^ In addition, cases are reported by OPs who tend to deal mostly with public sector workers, and therefore are less likely to represent private sector workers,^28^ with potentially different precipitating events for WRS.^29^ Therefore, any changes in trend would also need to consider which sector the cases were primarily derived from.

In summary, new cases of WRS are still most associated with the domains *demands* and *relationships* experienced at work. The trends in any new cases associated with these two domains have been gradually declining since introduction of the MS in 2004. Secondary additional precipitating events, (usually *change, support, control*, and *role)* are creating more complex cases with two or more precipitating events that appear to be more common over time. This analysis does not provide definitive evidence for the influence of the MS tool, further work is still required to unpick the reasons why changes have occurred, such as differences in underlying diagnoses (i.e. more specifically anxiety, depression, and stress), and occupational and industry differences. Given HSE’s renewed focus on WRMIH post-COVID, with focus on changing working environments since 2020^30-33^. The work here therefore adds to our understanding of the MS tool and its usefulness with respect to WRS.

## Data Availability

All data produced in the present study are only available upon reasonable request to the THOR team.
https://sites.manchester.ac.uk/thor/

## Notes

**COI** DF is an Honorary consultant respiratory physician Manchester University NHS Foundation Trust, and Chief Medical Advisor to the HSE. Though the work was partly funded by the HSE support for the THOR scheme, the work was devised, designed, and carried out entirely independently of the HSE. The contents of this publication are the views of the authors, and not necessarily those of HSE or HSE policy makers. Otherwise, the author team has no known conflict of interests.

### Competing Interest Statement

DF is an Honorary consultant respiratory physician Manchester University NHS Foundation Trust, and Chief Medical Advisor to the HSE. Though the work was partly funded by the HSE support for the THOR scheme, the work was devised, designed, and carried out entirely independently of the HSE. The contents of this publication are the views of the authors, and not necessarily those of HSE or HSE policy makers. Otherwise, the author team has no known conflict of interests.

### Funding Statement

Funding for THOR is provided by the HSE. This paper however was completed independently and only expresses the views of the authors and not necessarily the funding body

### Author Declarations

THOR has National Health Service ethics approval given by the North-West (Haydock) Research Ethics Committee (reference 17/NW/0129).

