## Supplementary Material for "Evaluating trends in new cases of Work-Related Mental Ill-health after introduction of The Health and Safety Executives’ Management Standards"

**Supplementary materials**

**Table S1; Aggregated combinations of Management Standards as reported as precipitating events to a case of work-related stress**

| Aggregated Combination | Freq | Percentage |
|---|---|---|
| *Not Matched to MS* | 2614 | 24.2 |
| *Demand Only* | 2570 | 23.8 |
| *Relationships Only* | 2546 | 23.5 |
| *Change Only* | 821 | 7.59 |
| *Support Only* | 439 | 4.06 |
| Demand+Change | 342 | 3.16 |
| Demand+Relationships | 255 | 2.36 |
| Demand+Support | 204 | 1.89 |
| *Control Only* | 156 | 1.44 |
| Relationships+Change | 112 | 1.04 |
| Relationships+Support | 103 | 0.95 |
| *Role Only* | 67 | 0.62 |
| Demand+Control | 65 | 0.60 |
| Demand+Role | 60 | 0.55 |
| Change+Support | 60 | 0.55 |
| Demand+Relationships+Change | 49 | 0.45 |
| Demand+Support+Control | 41 | 0.38 |
| Demand+Relationships+Support | 36 | 0.33 |
| Demand+Change+Support | 35 | 0.32 |
| Relationships+Role | 22 | 0.20 |
| Support+Role | 20 | 0.18 |
| Change+Role | 18 | 0.17 |
| Demand+Relationships+Control | 17 | 0.16 |
| Relationships+Control | 17 | 0.16 |
| Demand+Support+Role | 11 | 0.10 |
| Demand+Relationships+Support+Control | 11 | 0.10 |
| Relationships+Change+Support | 10 | 0.09 |
| Demand+Change+Control | 10 | 0.09 |
| Demand+Control+Role | 10 | 0.09 |
| Relationships+Support+Control | 9 | 0.08 |
| Change+Control | 9 | 0.08 |
| Demand+Relationships+Role | 8 | 0.07 |
| Demand+Change+Role | 8 | 0.07 |
| Relationships+Change+Role | 7 | 0.06 |
| Support+Control | 7 | 0.06 |
| Change+Support+Role | 6 | 0.06 |
| Change+Support+Control | 6 | 0.06 |
| Demand+Change+Control+Role | 4 | 0.04 |
| Demand+Change+Support+Role | 4 | 0.04 |
| Relationships+Change+Control | 4 | 0.04 |
| Demand+Change+Support+Control | 3 | 0.03 |
| Control+Role | 3 | 0.03 |
| Change+Control+Role | 2 | 0.02 |
| Relationships+Control+Role | 2 | 0.02 |

| | | |
|---|---|---|
| Demand+Relationships+Control+Role | 2 | 0.02 |
| Demand+Relationships+Support+Role | 2 | 0.02 |
| Relationships+Change+Support+Role | 2 | 0.02 |
| Demand+Relationships+Change+Control | 1 | 0.01 |
| Relationships+Change+Control+Role | 1 | 0.01 |
| Change+Support+Control+Role | 1 | 0.01 |
| Demand+Change+Support+Control+Role | 1 | 0.01 |
| Demand+Relationships+Change+Support+Control+Role | 1 | 0.01 |
| Relationships+Support+Role | 1 | 0.01 |
| **Total** | **10815** | |
| **Single Management Standard identified** | 6599 | 61.0 |
| **Two Management Standards identified** | 1297 | 12.0 |
| **Three Management Standards identified** | 272 | 2.5 |
| **Four or more Management Standards identified** | 33 | 0.3 |
| **No Management Standard identified** | 2614 | 24.2 |

**Table S2: Frequency of monthly counts between 1996-2019 of work-related stress by any reported occurrence and the aggregated Management Standards groups.**

| Management Standards | No. Cases of work-related stress (Row%) reported per month | | | | | |
|---|---|---|---|---|---|---|
| (Combinations of occurrence) | 1 | 2-3 | 4-5 | 6-10 | >10 | Total months (col%) |
| WRS Not assigned MS | 1062(66.0) | 452(28.1) | 68(4.2) | 25(1.6) | 3(0.2) | 1610(22.0) |
| Demands Only | 1138(68.1) | 457(27.3) | 62(3.7) | 13(0.8) | 2(0.1) | 1672(22.8) |
| Relationships Only | 1072(65.2) | 500(30.4) | 59(3.6) | 12(0.7) | 1(0.1) | 1644(22.4) |
| Change Only | 518(80.1) | 120(18.5) | 8(1.2) | 1(0.2) | - | 647(8.8) |
| Support Only | 296(83.1) | 57(16.0) | 3(0.8) | - | - | 356(4.9) |
| Demands + Other | 636(75.2) | 184(21.7) | 24(2.8) | 2(0.2) | - | 846(11.6) |
| Relationships + Other | 220(86.6) | 34(13.4) | - | - | - | 254(3.5) |
| Any Other combination | 251(85.4) | 39(13.3) | 3(1.0) | 1(0.3) | - | 294(4.0) |
| **Management Standards (Any)** | **1** | **2-3** | **4-5** | **6-10** | **>10** | **Total Months (col%)** |
| Assigned a MS | 4131(72.3) | 1391(24.3) | 159(2.8) | 29(0.5) | 3(0.1) | 5713(78.0) |
| Not Assigned a MS | 1062(66.0) | 452(28.1) | 68(4.2) | 25(1.6) | 3(0.2) | 1610(22.0) |
| WRS Not assigned Demands | 3419(71.2) | 1202(25.0) | 141(2.9) | 39(0.8) | 4(0.1) | 4805(65.6) |
| Demands | 1774(70.5) | 641(25.5) | 86(3.4) | 15(0.6) | 2(0.1) | 2518(34.4) |
| WRS Not assigned Relationships | 3680(72.4) | 1212(23.8) | 148(2.9) | 41(0.8) | 5(0.1) | 5086(69.5) |
| Relationships | 1513(67.6) | 631(28.2) | 79(3.5) | 13(0.6) | 1(0.0) | 2237(30.5) |
| WRS Not assigned Change | 4226(69.7) | 1581(26.1) | 200(3.3) | 51(0.8) | 6(0.1) | 6064(82.8) |
| Change | 967(76.8) | 262(20.8) | 27(2.1) | 3(0.2) | - | 1259(17.2) |
| WRS Not assigned Support | 4562(70.4) | 1654(25.5) | 206(3.2) | 53(0.8) | 6(0.1) | 6481(88.5) |
| Support | 631(74.9) | 189(22.4) | 21(2.5) | 1(0.1) | - | 842(11.5) |
| WRS Not assigned Control | 4983(71.0) | 1758(25.1) | 214(3.1) | 53(0.8) | 6(0.1) | 7014(95.8) |
| Control | 210(68.0) | 85(27.5) | 13(4.2) | 1(0.3) | - | 309(4.2) |
| WRS Not assigned Role | 5017(70.9) | 1784(25.2) | 220(3.1) | 54(0.8) | 6(0.1) | 7081(96.7) |
| Role | 176(72.7) | 59(24.4) | 7(2.9) | - | - | 242(3.3) |
| **Total** | **4131(56.4)** | **1391(19.0)** | **159(2.2)** | **29(0.4)** | **3(0.0)** | **7,323** |

**Figure S1:** Annual incidence rate ratios (IRRs) with 95% confidence intervals referenced against 2019 for work-related stress for aggregated combinations of mentions of Management Standards (implemented in 2004).

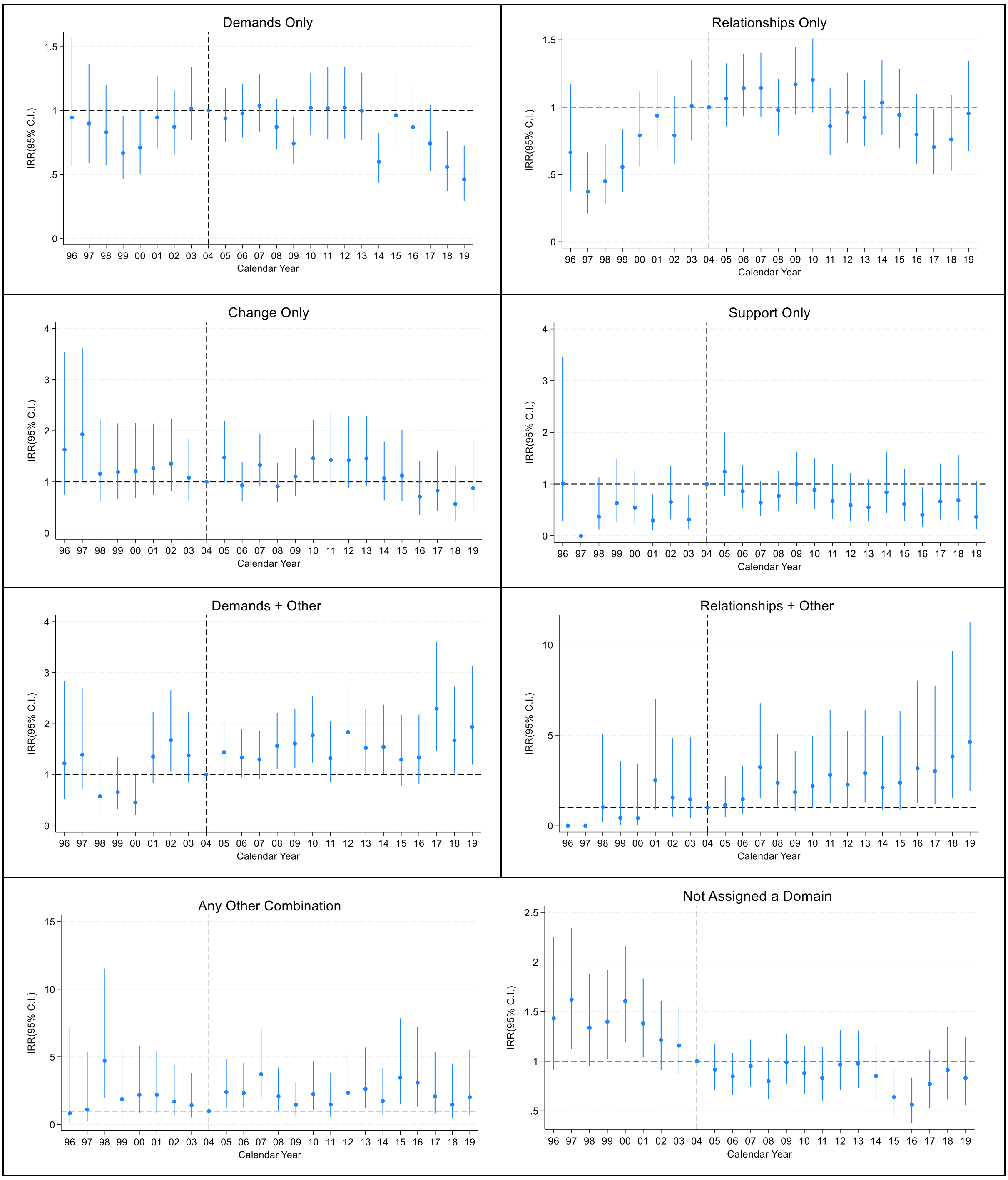

**Figure S2:** Annual incidence rate ratios (IRRs) with 95% confidence intervals referenced against 2019 for work-related stress for 'any' mention of Management Standards (implemented in 2004).

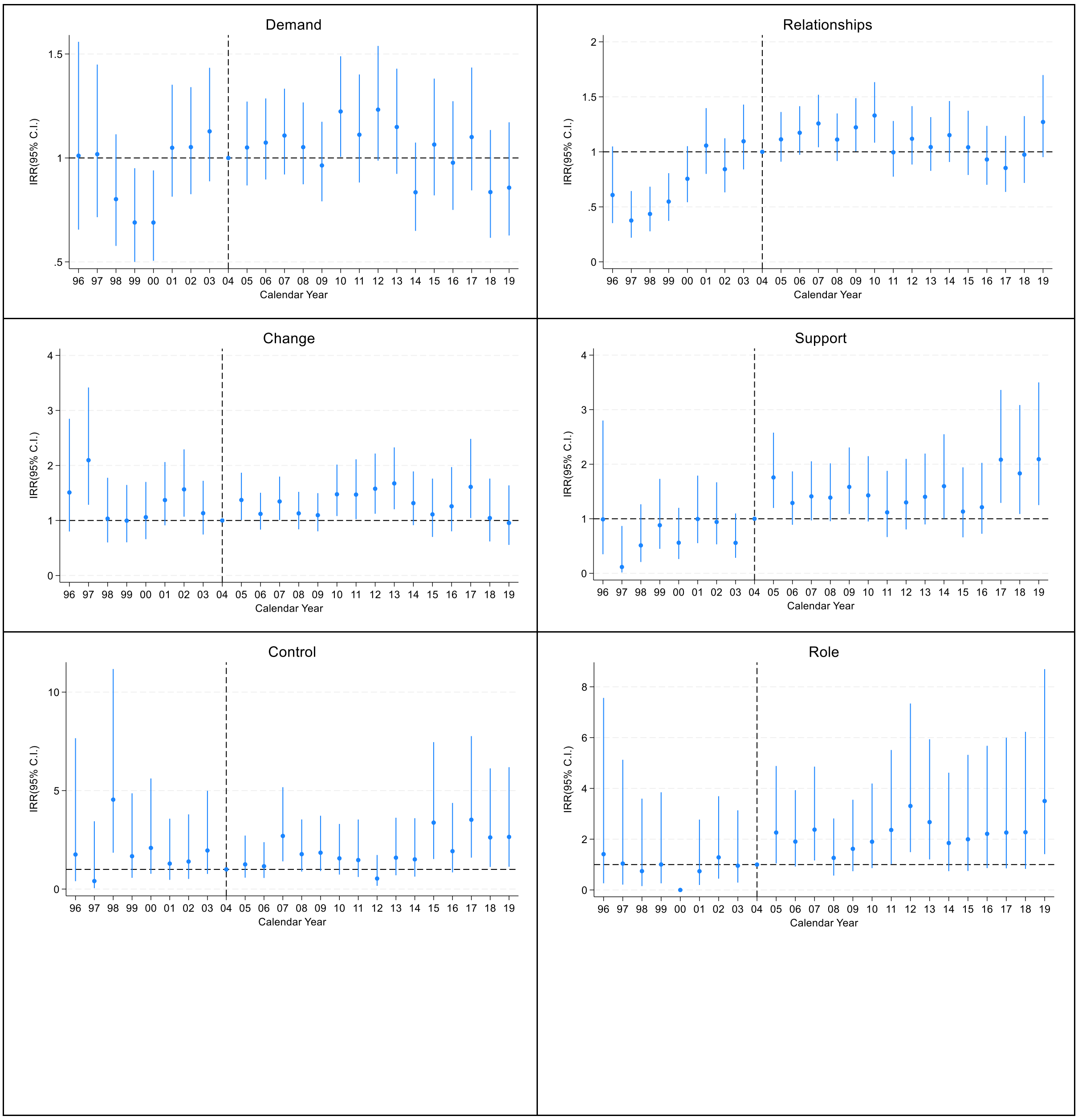

**Figure S3:** Annual stacked count and percentages of new cases of work-related stress split by aggregated groups of the associated Management Standards

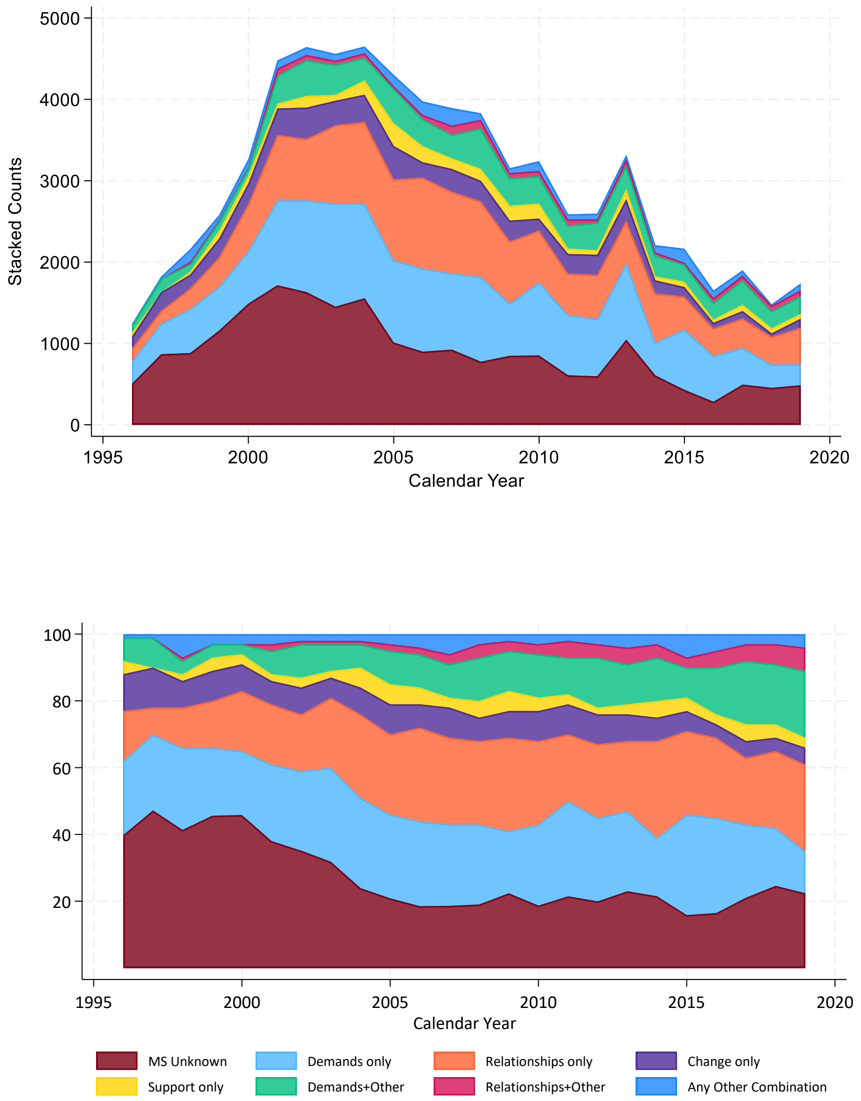

**Table S3** – Sensitivity analysis time-interrupted cut point set to be 2006 (end of Management Standard awareness period) and 2004 (implementation of Management Standards): Annual percentage change IRR (95%C.I.) comparing 1996-2019 trends in Management Standards mentioned in association with the Work-Related Stress (WRS) monthly counts of cases

| Trends (1996-2019)[a] | Total WRS[1] | Demand[2] | Relationships[2] | Change[2] | Support[2] | Control[2] | Role[2] |
|---|---|---|---|---|---|---|---|
| **Trends (split at 2006)** | | | | | | | |
| **Calendar Year(pre2006)** | 1.23 (-0.01,2.56) | 3.46 (1.12,5.85) | 9.78 (6.90,12.7) | -0.84 (-4.32,2.78) | 11.1 (5.37,17.2) | -3.40 (-10.5,4.27) | 13.3 (2.63,25.1) |
| **Calendar Year(2006-)** | -1.31 (-2.07,-0.50) | -0.97 (-2.24,0.32) | -1.60 (-2.93,-0.25) | 1.08 (-0.98,3.18) | 1.39 (-1.09,3.94) | 4.88 (0.70,9.24) | 3.29 (-1.14,7.93) |
| **Random Effect Variance** | 0.13 | 0.25 | 0.36 | 0.51 | 0.74 | 1.35 | 1.19 |
| **AIC** | 15,120 | 10,416 | 9,250 | 6,205 | 4,660 | 2,366 | 1,840 |
| | **Demands Only[3]** | **Relationships Only[3]** | **Change Only[3]** | **Support Only[3]** | **Demands + Other[3]** | **Relationships + Other[3]** | **Any Other Combinations[3]** |
| **Calendar Year(pre2006)** | 2.83 (0.12,5.62) | 8.99 (5.91,12.2) | -1.65 (-5.95,2.85) | 9.33 (1.98,17.2) | 5.82 (1.16,10.7) | 17.0 (4.71,30.7) | 2.34 (-4.96,10.2) |
| **Calendar Year(2006-)** | -2.75 (-4.28,-1.21) | -2.85 (-4.32,-1.36) | -1.18 (-3.90,1.62) | -4.86 (-8.33,-1.27) | 2.08 (-0.23,4.44) | 4.81 (0.63,9.16) | 0.05 (-3.92,4.19) |
| **Random Effect Variance** | 0.33 | 0.38 | 0.53 | 0.96 | 0.83 | 1.34 | 0.87 |
| **AIC** | 8,512 | 8,244 | 4,290 | 2,670 | 5,054 | 1,811 | 2,388 |

1. Trends analysis of all WRS cases
2. Trends analysis of WRS due to 'any' mention of management standards category (e.g. Count of any mention of control)
3. Trends analysis of WRS due to 'common combinations' management standards mentioned in association with the WRS (i.e. more Demands and Relationships may be associated with the WRS case)

a = Adjusted for calendar month, first month as reporter, and 'core' vs 'sample' reporter
